# Appearance of IgG to SARS-CoV-2 in saliva effectively indicates seroconversion in mRNA vaccinated immunocompromised individuals

**DOI:** 10.1101/2021.09.30.21264377

**Authors:** Katie Healy, Elisa Pin, Puran Chen, Gunnar Söderdahl, Piotr Nowak, Stephan Mielke, Lotta Hansson, Peter Bergman, C. I. Edvard Smith, Per Ljungman, Davide Valentini, Ola Blennow, Anders Österborg, Giorgio Gabarrini, Khaled Al-Manei, Hassan Alkharaan, Michal Jacek Sobkowiak, Xinling Xu, Mira Akber, Karin Loré, Cecilia Hellström, Sandra Muschiol, Gordana Bogdanovic, Marcus Buggert, Hans-Gustaf Ljunggren, Sophia Hober, Peter Nilsson, Soo Aleman, Margaret Sällberg Chen

**Author notes:** **Corresponding authors:** Margaret Sällberg Chen, DDS, PhD, Professor, Dept. of Dental Medicine, Karolinska Institutet, 141 04 Huddinge, Sweden, MAIL, http://ki.se/people/marche; Soo Aleman, MD, PhD, Assoc Professor, Head of HIV, Viral hepatitis and Immunodeficiency disorders units, Department of Infectious Diseases, Karolinska University Hospital, Sweden, MAIL. KH, EP and PC shared first authorship. SA and MSC shared senior authorship. **AUTHOR CONTRIBUTIONS** SA, EP, HGL, PN, MSC conceived the project. KH, PC, GG, KAM, HA, MJS, MA, MSC established the protocols for handling clinical material. GS, PN, SM, LH, PB, CIES, PL, XX, OB, AÖ collected and compiled clinical data. EP, CH, SM, GB performed measurements and analyzed the laboratory data. KH, PC and DV performed statistics. SA, EP, SH, PN, HGL, MSC provided resources and supervised the project. KH, EP, SA and MSC wrote the original draft. All reviewed and edited the manuscript.

## Abstract

**Background:** Immunocompromised individuals are highly susceptible to severe acute respiratory syndrome coronavirus-2 (SARS-CoV-2) infection. Whether vaccine-induced immunity in these individuals involves the oral cavity, a primary site of infection, is presently unknown.

**Methods:** Immunocompromised individuals (n=404) and healthy controls (n=82) participated in a prospective clinical trial encompassing two doses of the mRNA BNT162b2 vaccine. Immunocompromised individuals included primary immunodeficiencies (PID) and secondary immunodeficiencies caused by human immunodeficiency virus (HIV) infection, allogeneic hematopoietic stem cell transplantation (HSCT)/chimeric antigen receptor T cell therapy (CAR-T), solid organ transplantation (SOT), and chronic lymphocytic leukemia (CLL). Saliva and serum samples were collected at four time points from the first vaccine dose until 2 weeks after second dose. SARS-CoV-2 spike specific immunoglobulin G (IgG) responses were quantified by a multiplex bead-based assay in saliva and correlated to paired serum IgG titers determined by Elecsys® Anti-SARS-CoV-2 S assay.

**Results:** IgG responses to the SARS-CoV-2 spike full-length trimeric glycoprotein (Spike-f) and S1 subunit in saliva in the HIV and HSCT/CAR-T groups were comparable to healthy controls. In contrast, PID, SOT, and CLL patients all displayed weaker responses which were mainly influenced by disease parameters or immunosuppressants. Salivary IgG levels strongly correlated with serum IgG titers on days 21 and 35 (rho=0.8079 and 0.7768, p=<0.0001). Receiver operating characteristic curve analysis for the predictive power of salivary IgG yielded AUC=0.95, PPV=90.7% for the entire cohort on D35.

**Conclusions:** Saliva conveys humoral responses induced by BNT162b2 vaccination. The predictive power makes it highly suitable for screening low responding/vulnerable groups for revaccination.

**Trial Registration:** ClinicalTrials.gov Identifier: NCT04780659

**Funding:** Knut and Alice Wallenberg Foundation, Erling Perssons family foundation, Region Stockholm, Swedish Research Council, Karolinska Institutet, The Swedish Blood Cancer Foundation and the organization for PID patient group in Sweden, and Nordstjernan AB. Center for Medical Innovation (CIMED), the Swedish Medical Research Council and the Stockholm County Council (ALF).

**GRAPHIC ABSTRACT:** 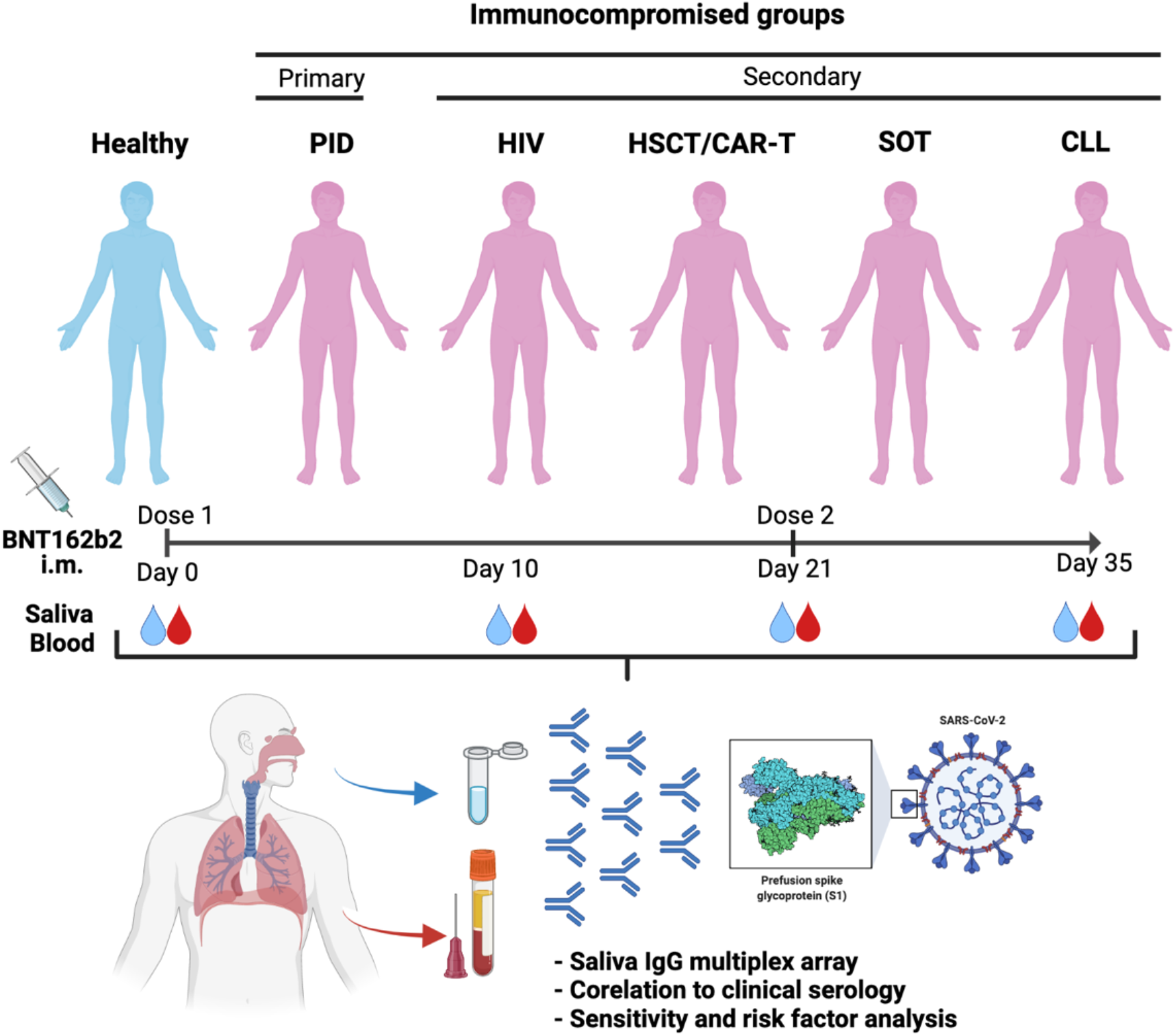

## INTRODUCTION

Vaccine development has been a success story of the coronavirus disease 2019 (COVID-19) pandemic. Among approved vaccines, the BNT162b2 vaccine (Comirnaty^®^, Pfizer-BioNTech) relies on novel mRNA technology, where mRNA is packaged into lipid nanoparticles to deliver genetic instructions for human cells to produce the SARS-CoV-2 spike protein (1). Accumulating data from the general population in Israel and early studies in US healthcare workers confirmed that vaccination with a two-dose regimen confers 94.6% and 95% protection against symptomatic infection and severe disease, respectively, 1-2 weeks after the second dose (2-4). In a more recent UK study, two doses were shown to be approximately 85-90% effective in adults aged 70 years and older (5). In contrast, data from studies in older adults receiving a single dose of BNT162b2 have yielded mixed results (6-8).

Adult patients with primary immunodeficiency (PID) or secondary immunodeficiency (SID) generally display higher morbidity and mortality rates from COVID-19 than immunocompetent individuals (9-11). The overall infection fatality rate (IFR) for PID and SID have been reported to be as high as 20% (PID) and 33% (SID), compared to less than 1% in the general population (9). Around six million people worldwide are estimated to live with a PID (12, 13), whilst SID disorders are frequent consequences of underlying medical conditions, e.g. human immunodeficiency virus (HIV) infection, malignant diseases, or clinical interventions with immunosuppressive drugs (14). Patients receiving immunosuppression after undergoing hematopoietic stem-cell transplantation (HSCT), specific cellular therapies (e.g. CAR-T cell therapy), or having hematological malignancies, often show prolonged virus shedding and transmission dynamics in which shedding of viable SARS-CoV-2 could be prolonged up to two months or more due to weakened immunity (15, 16). Notably, people with compromised immunity have been mostly excluded from large clinical trials addressing mRNA vaccine-effectiveness (2, 17). Recent published reports have however indicated weak or absent immune responses in several groups of immunocompromised persons (18-22).

Mucosal immunity in the aerodigestive tract is considered a front-line defense against SARS-CoV-2 infection. The oral cavity is an important site for SARS-CoV-2 infection and saliva is considered a potential route of virus transmission (23). Transmission can occur by activities involving the oral cavity, such as breathing, coughing, sneezing, speaking, or singing (24-26). Oral manifestations such as taste loss, dry mouth, and oral lesions are present in about half of confirmed COVID-19 cases (27). Viral entry factors such as ACE2 and TMPRSS2, TMPRSS4, and TMPRSS11D are expressed in the oral cavity (buccal mucosa, ventral tongue, and the dorsal tongue) and the oropharynx (soft palate and tonsils), including salivary glands and epithelial cells in saliva (23). It was recently shown that saliva antibodies correlate with seroconversion in mRNA vaccinated healthcare workers (15, 16). However, whether mRNA vaccines such as the BNT162b2 vaccine can induce mucosal immunity at distal sites, such as the oral cavity, following intramuscular injection in immunocompromised patients is presently unknown. Immunocompromised patients in this context represent a large and highly important risk group in need of continuous monitoring of vaccination efficacy.

To fill the knowledge gap in respect to COVID-19 vaccine efficacy, we recently conducted a prospective open-label clinical trial (COVAXID, EudraCT, no. 2021-000175-37) investigating the immunogenicity of the BNT162b2 vaccine in immunocompromised patients and healthy controls (28). The aim of the present study was to investigate vaccine-induced humoral immunity in the oral cavity in the same cohort.

## RESULTS

### Study design and patient demographics

From the COVAXID clinical trial (539 participants; 449 patients and 90 controls), 486 participants, 404 immunocompromised and 82 healthy participants were eligible for inclusion in the present study. Patient parameters are shown in **Table 1**. As presented in the accompanying flow chart (**Supplementary figure 1**), eligible participants had to be SARS-CoV-2 seronegative at baseline and not meet exclusion criteria such as PCR-positivity at any point of the study, missing baseline serum antibody data, or fewer than two vaccine doses. Saliva and serum samples were collected at four time points; days 0 (D0), 10 (D10), 21 (D21) and 35 (D35) from first vaccine dose. The second vaccine dose was administered at D21. A total of 1870 saliva samples were obtained with 1829 paired serum sampled across all timepoints. The saliva flow rate in most participants was above normal (>0.1 mL/min) at each time point measured, albeit a lower mean flow rate was seen in the PID (p=0.0392) and CLL groups (p<0.0001) (**Table 1 and Supplementary figure 2**).

**Table 1.** Baseline characteristics of study participants and blood and saliva status.

| Group | Healthy controls | All patients | PID | HIV | HSCT/<br>CAR-T | SOT | CLL |
| --- | --- | --- | --- | --- | --- | --- | --- |
| **n after exclusion** | 82 | 404 | 79 | 80 | 74 | 83 | 88 |
| **Sex, Men (%):**<br>**Women (%)** | 35 (43%):<br>47 (57%) | 217 (54%):<br>186 (46%) | 30 (38%):<br>49 (62%) | 47 (59%):<br>33 (41%) | 40 (54%):<br>33 (45%) | 42 (51%):<br>41 (49%) | 58 (66%):<br>30 (34%) |
| **Age (years)**<br>**[median, IQR]** | 54<br>(34-67) | 60<br>(47-69) | 50<br>(37-59) | 54<br>(42-63) | 60<br>(51-67) | 59<br>(47-67) | 70<br>(62-76) |
| **Blood**<br>**parameters§**<br>**[median, IQR]** |  |  |  |  |  |  |  |
| IgG (g/L) | 11 (10-13) | 10 (9-12) | 13 (10-15) | 10 (7-12) | 10 (7-11) | 6.50 (5-10) | 10 (9-12) |
| Absolute<br>lymphocyte count<br>(x10<sup>9</sup>/L) | 1.60<br>(1.10-2.40) | 1.3<br>(0.9-1.60) | 1.8<br>(1.4-2.30) | 1.6<br>(1-2.70) | 1.3<br>(0.88-1.70) | 5.9<br>(1.6-22) | 1.60<br>(1.10-2.40) |
| Creatinine (μmol/L) | 70.50<br>(63-81) | 80<br>(65-95) | 66<br>(57-79) | 83.5<br>(67-99) | 79<br>(62-91) | 98<br>(79-119) | 80<br>(67-88) |
| **Ongoing**<br>**immunosuppression,**<br>**n (%)** |  |  |  |  |  |  |  |
| Corticosteroids | 0 (0%) | 94 (23%) | 9 (11%) | 0 (0%) | 9 (12%) | 76 (92%) | 0 (0%) |
| Other<br>immunosuppressive<br>agents | 0 (0%) | 144 (36%) | 12 (15%) | 0 (0%) | 18 (26%) | 83 (100%) | 30 (34%) |
| **Systemic IgG**<br>**substitution,**<br>**n (%)** | 0 (0%) | 51 (13%) | 51 (65%) | 0 (0%) | 0 (0%) | 0 (0%) | 0 (0%) |
| **Saliva flow rate**<br>**(mL/min) [median,**<br>**IQR] §§** |  |  |  |  |  |  |  |
| D0 | 0.59<br>(0.4-0.7) | 0.56<br>(0.4-0.7) | 0.54<br>(0.4-0.7) | 0.55<br>(0.4-0.7) | 0.66<br>(0.5-0.8) | 0.56<br>(0.4-0.8) | 0.46<br>(0.2-0.7) |
| D10 | 0.60<br>(0.4-0.8) | 0.62<br>(0.4-0.8) | 0.60<br>(0.3-0.8) | 0.64<br>(0.4-0.8) | 0.63<br>(0.4-0.8) | 0.68<br>(0.5-0.8) | 0.59<br>(0.4-0.8) |
| D21 | 0.60<br>(0.5-0.7) | 0.58<br>(0.4-0.7) | 0.55<br>(0.4-0.7) | 0.60<br>(0.4-0.8) | 0.66<br>(0.5-0.8) | 0.60<br>(0.4-0.7) | 0.45<br>(0.3-0.7) |
| D35 | 0.66<br>(0.4-0.8) | 0.60<br>(0.4-0.8) | 0.54<br>(0.3-0.8) | 0.62<br>(0.5-0.8) | 0.62<br>(0.4-0.8) | 0.62<br>(0.4-0.8) | 0.53<br>(0.3-0.7) |

§Measured at treatment initiation
§§ Number of days after vaccine dose 1.
Abbreviations: n = number; PID = primary immunodeficiency disorders; HIV = human immunodeficiency virus; HSCT = hematopoietic stem cell transplantation; SOT = solid organ transplantation; CLL = chronic lymphocytic leukemia; IgG = immunoglobulin G; CAR-T = chimeric antigen receptor T-cell therapy.

### Anti-spike IgG responses in saliva are related to immunodeficiency status

All groups showed a steady induction of anti-spike IgG reactivities in saliva after the first vaccine dose, where people living with HIV, hereafter referred to as “HIV”, and healthy controls exhibited the earliest and largest increase relative to baseline (D0) (**Figure 1A**). From baseline to D21 (before second dose), the IgG reactivates to Spike-f (full-length spike, trimeric form stabilized in prefusion-conformation) increased by 12- and 9-fold in healthy controls and the HIV group, respectively; thereafter to 74- and 53-fold after the second dose (D35). In these groups, most participants (>90%) developed anti-spike IgG (both Spike-f and S-1) in saliva at D35. In the HSCT/CAR-T group, a moderate 3-fold increase in salivary Spike-f IgG reactivity was observed at D21. After the second dose it rose to 50-fold relative to baseline by D35 indicating a potent response after full vaccination. In contrast, weak responses were seen in the PID, SOT, and CLL groups in which a discrete increase (1-2-fold) in D21 samples was found relative to baseline. Not until D35 did a sizeable fraction of PID, SOT, and CLL patients demonstrate a detectable anti-Spike-f IgG reactivity in saliva with median values of 4-5 fold over baseline in PID and CLL groups and less in SOT group, albeit many patients in these three groups remained negative. It was noted that Spike S1-specific IgG reactivities were similar as seen for the Spike-f antigen.

**Figure 1:**
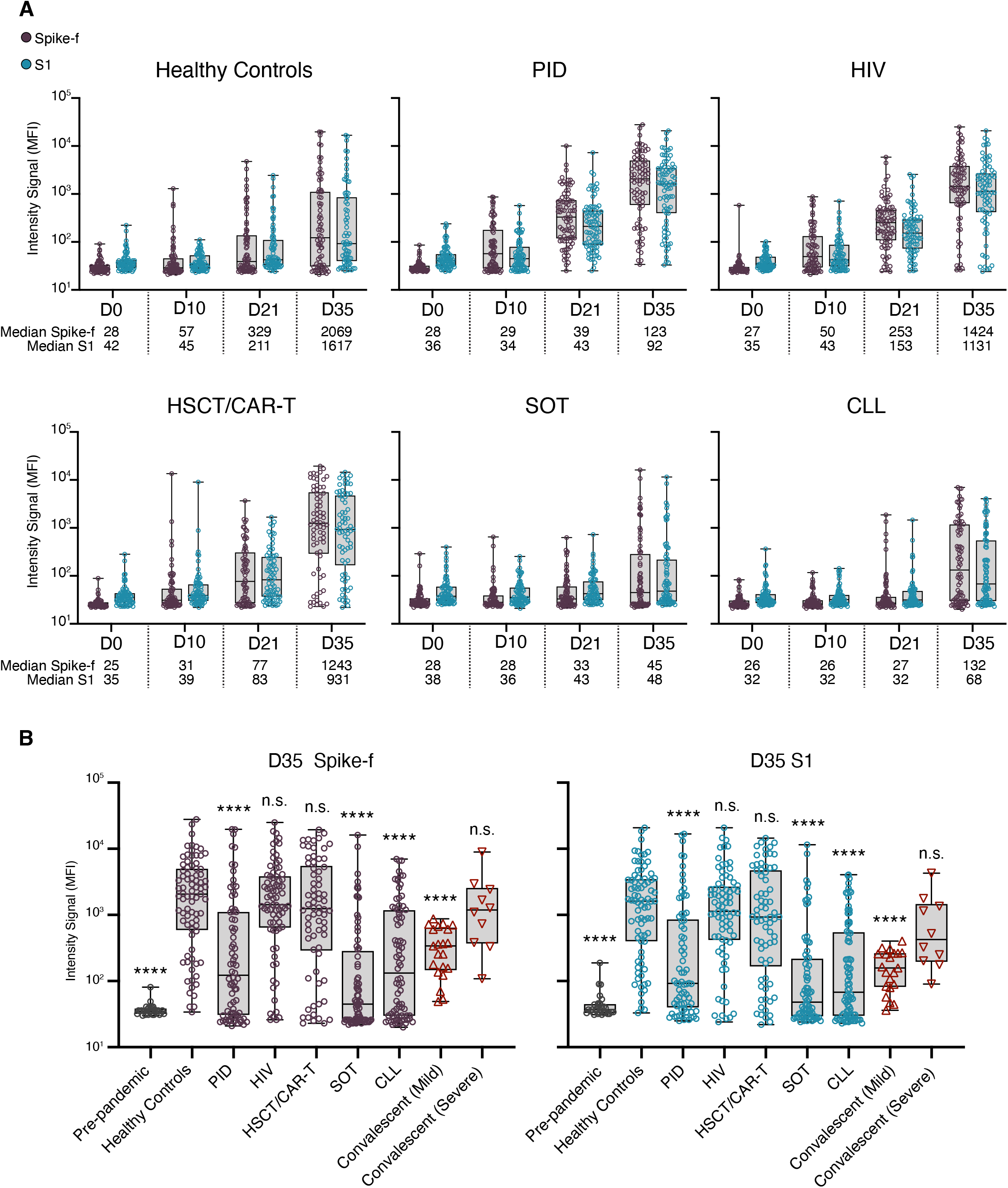
SARS-CoV-2 spike-specific IgG responses in saliva in immunocompetent or immunocompromised individuals. (A) Levels of salivary Spike-f and S1 IgG responses on days 0, 10, 21, and 35 after first vaccine dose. Healthy controls: D0; D10; D21; D35 (n=82; n=77; n=81; n=81), PID: D0; D10; D21; D35 (n=78; n=77; n=78; n=74), HIV: D0; D10; D21; D35 (n=79; n=78; n=77; n=77), HSCT/CAR-T: D0; D10; D21; D35 (n=73; n= 74; n=73; n=70), SOT: D0; D10; D21; D35 (n=78; n=80; n=80; n=76), CLL: D0; D10; D21; D35 (n=85; n=78; n=82; n=82) (B) Comparison of D35 Spike-f and S1 IgG responses in saliva of vaccinated healthy controls relative to indicated patient group or control non-vaccinated groups (pre-pandemic (n=29) and respective mild/severe (n=21/n=10) convalescent individuals). Lines, boxes, and whiskers represent the median, IQR, and min-max range, respectively. The Mann-Whitney U test was used for group comparisons against healthy controls in (B). **** p<0.0001. ns = not significant.

A summary of all saliva data collected at D35, i.e., 14 days after the second vaccine dose from all groups were compared, using COVID-19 convalescence saliva and pre-pandemic saliva samples also as references (**Figure 1B**). Among immunocompromised patient groups, the strongest magnitude of anti-Spike-f and anti-S1 responses in saliva was observed in the HIV and HSCT/CAR-T groups at D35, which did not differ from the healthy controls. In contrast, the PID, SOT, and CLL patient groups all had lower SARS-CoV-2 specific responses in saliva on D35 relative to healthy controls (p<0.001). Additionally, salivary IgG to both spike antigens in the healthy controls, HIV and HSCT/CAR-T groups were higher than convalescence saliva collected from mild COVID-19 patients (p<0.001) and were of similar magnitude as the severe COVID-19 convalescent saliva.

### SARS-CoV-2 specific IgG responses in saliva and serum strongly correlate

To evaluate whether SARS-CoV-2 specific IgG responses in saliva corresponded to those in serum, paired analyses were performed across all timepoints. Serum anti-S1 antibody data was generated using the quantitative test Elecsys^®^ Anti-SARS-CoV-2 S (29) that has been validated on serum samples from patients and against the WHO reference standard (WHO/BS/2020.2402). To assess saliva as a diagnostic indicator of serum responses, Spearman correlation analysis in paired samples was performed for the entire cohort at D10 (n=445), D21 (n=464), and D35 (n=445). As shown in **Figure 2**, a moderate correlation was observed by D10 (r=0.4795, p<0.001), followed by strong correlations at D21 (r=0.8079, p<0.001) and D35 (r=0.7768, p<0.001). Similar temporal correlations were also found between anti-S1 IgG in saliva and paired serum spike RBD IgG levels (**Supplementary figure 3**). Correlating the D35 salivary Spike-f or S1 IgG reactivities on a group level to serum anti-S1 antibody titers demonstrated moderate correlations in the healthy controls (rho=0.4290 and 0.4303, p<0.0001) and HIV (rho=0.3886 and 0.4025, p<0.001) groups, and strong correlations in the PID (rho=0.8500 and 0.8247, p<0.0001), HSCT/CAR-T (rho=0.8331 and 0.8359, p<0.0001), SOT (rho=0.7582, p<0.0001) and CLL (rho=0.6951 and 0.6660, p<0.0001) groups (**Table 2**). Taken together, this data confirmed there is a strong agreement between the salivary spike IgG (irrespective of Spike-f or S1) and serum spike IgG with the latter measured by an independent clinical laboratory. This concordance was seen at the cohort level as well as patient group level particularly after day 21 from first vaccine dose.

**Table 2.** Correlations between salivary spike IgG MFI and serum IgG spike S1 RBD IgG IU/ml in D35 saliva samples.

| Ab specificity | Spike-f |  | S1 |  |
| --- | --- | --- | --- | --- |
|  | Spearman rho | p-value | Spearman rho | p-value |
| **Healthy Controls** | 0.4290 | <0.0001 | 0.4303 | <0.0001 |
| **PID** | 0.8500 | <0.0001 | 0.8247 | <0.0001 |
| **HIV** | 0.3886 | 0.0006 | 0.4025 | 0.0004 |
| **HSCT/CAR-T** | 0.8331 | <0.0001 | 0.8359 | <0.0001 |
| **SOT** | 0.7582 | <0.0001 | 0.6660 | <0.0001 |
| **CLL** | 0.6951 | <0.0001 | 0.7344 | <0.0001 |

Abbreviations: n = number, PID: primary immunodeficiency disorders, HIV: human immunodeficiency virus, HSCT: hematopoietic stem cell transplantation, SOT: solid organ transplantation, CLL: chronic lymphocytic leukemia, IgG: immunoglobulin G, CAR-T: chimeric antigen receptor T-cell therapy. Spearman correlation analysis was performed.

**Figure 2:**
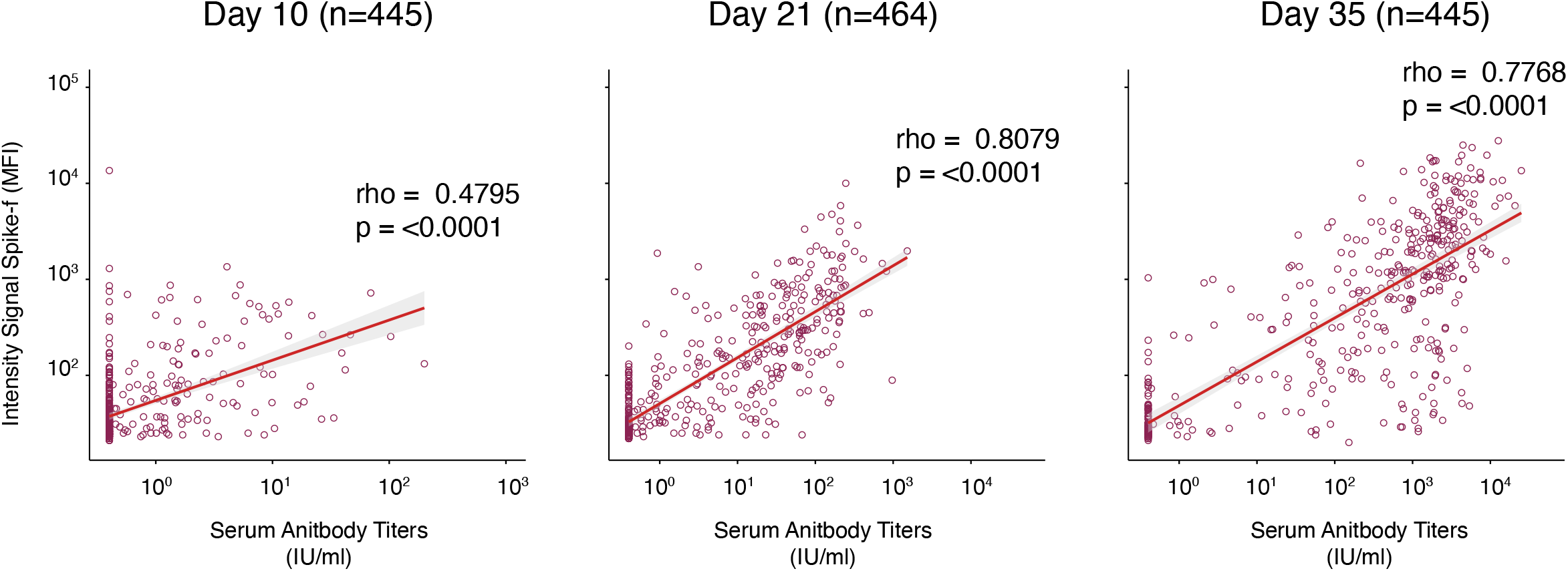
Correlation between spike IgG in paired serum and saliva. Salivary Spike-f IgG MFI signal intensity (y-axis) was measured by a multiplex bead-based assay, and serum SARS-CoV-2 spike IgG levels expressed as international units (x-axis) were measured by the quantitative test Elecsys® Anti-SARS-CoV-2 S. Correlation plots of the entire COVAXID cohort at D10, D21, and D35 post-vaccination. MFI = Median Fluorescence Intensity; IU = International Units. Spearman correlation analysis was used to determine rho- and p-values.

### Sex and age-based influences on SARS-CoV-2 specific responses in saliva and blood

To evaluate whether sex and age impacted SARS-CoV-2 vaccine responses in the study cohort, these parameters were analyzed on a group level in paired saliva and serum samples collected two weeks after the second dose (D35). Interestingly, a significant sex-based difference was observed in both serum and saliva in the PID group, with women demonstrating significantly stronger responses, while none of the other groups showed any significant sex-based influence (**Figure 3A**). Subgrouping the patient cohort by age (< 60 years/ ≥ 60 years) did not reveal any significant differences in saliva. However, a stronger serum SARS-CoV-2 specific IgG magnitude was found in the younger subgroup (<60 years) of healthy controls (**Figure 3B**). Taken together, except for the PID-group, sex and age appeared to have little impact on the saliva results.

**Figure 3:**
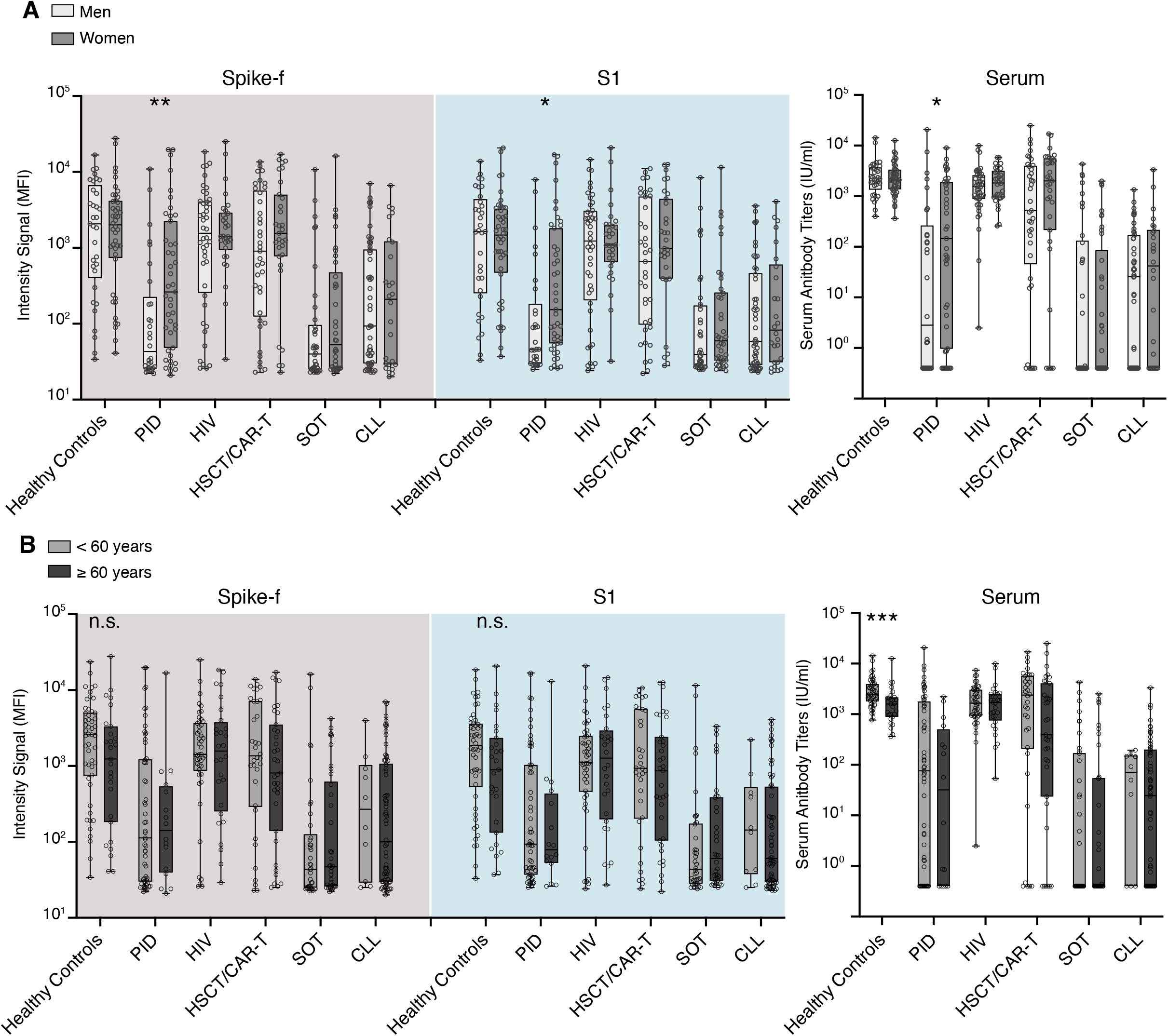
Sex and age have minimal impact on antibody responses detected in saliva and serum. (A) Sex and (B) age-based comparisons of salivary IgG to Spike-f and S1 MFI and serum IgG to spike of paired D35 saliva and serum samples from fully vaccinated individuals. MFI = Median Fluorescence Intensity; IU = International Units. Lines, boxes, and whiskers represent the median, IQR, and min-max range, respectively. The Mann-Whitney U test was used to test significance. **** p<0.0001, *** p<0.0002, ** p<0.0021, * p<0.0332. ns = not significant.

### Influences of disease and treatment status observed in both serum and saliva

The influence of disease status or treatment regimens on vaccine-induced SARS-CoV-2-specific responses were further examined on a patient subgroup level in paired saliva and serum samples on D35. As shown in **Figure 4**, further age stratification revealed no difference among healthy participants. Among PID patients, the CVID and XLA subgroups (n=39 and n=4, respectively) showed the lowest median antibody response in both saliva and serum, whilst subgroups with monogenic PID disease (n=9), CD4-cytopenia (n=11), or other PID disorders (n=10) generated responses close to healthy control levels. On the other hand, participants living with HIV with either low (n=22) or high (n=52) CD4 T cell counts had a similar range of median antibody levels in both saliva and serum compared to healthy controls. In the HSCT/CAR-T group, the lowest responses were seen in those patients receiving CAR T treatment (n=2) and in those being in an early or intermediate phase (<6 months) post-HSCT transplantation (n=3 and n=11, respectively). The responses were, however, close to healthy levels in the subgroup in the late phase post-HSCT transplantation (n=53). SOT patients had the lowest overall antibody response in both serum and saliva, with a particularly poor response in patients receiving mycophenolate mofetil (MMF) as a part of their immunosuppression regimen (n=46), while patients without MMF (n=30, all vaccinated >6 months after transplantation) had a moderate vaccine response. In the CLL subgroups, lowest antibody responses were observed in those receiving ibrutinib treatment (n=26) followed by those being off ibrutinib treatment (n=8), a BTK inhibitor that suppresses B-cell signaling. Although the responses varied among the CLL subgroups, a significant proportion of indolent or previously chemoimmunotherapy-treated (Prev CD20 mAb based therapy) CLL patients produced antibody responses in both serum and saliva. Based on these observations, the striking similarities in the SARS-CoV-2 specific IgG profile in saliva and serum observed even at the subgroup levels strengthens the usefulness for saliva as an indicator for seroconversion, which was measured by the quantitative clinical serology assay.

**Figure 4:**
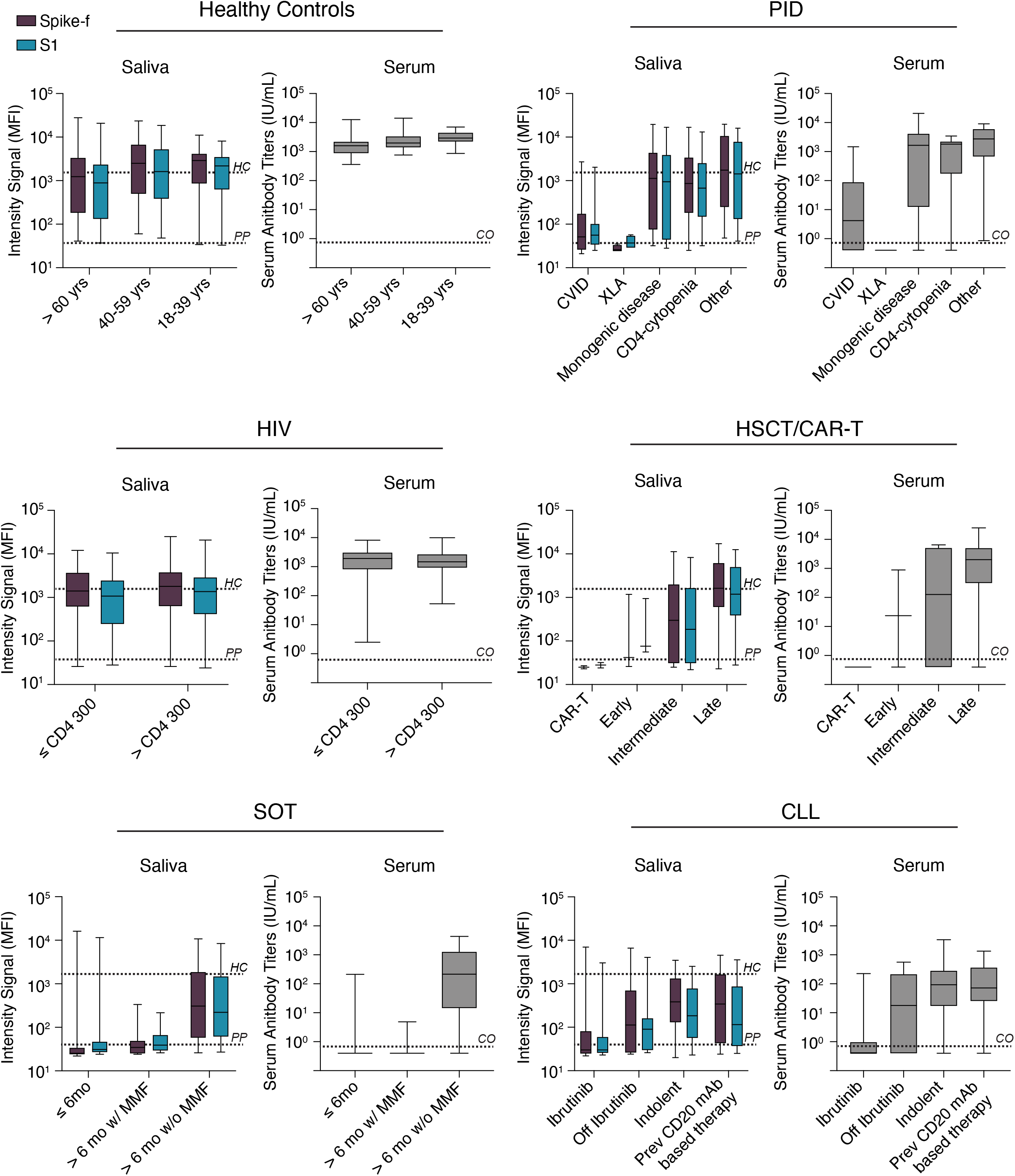
Patient subgroup analysis. (A) Disease or treatment status subgrouping of immunocompromised individuals included in the COVAXID study on D35 saliva or serum samples of fully vaccinated individuals. Lines, boxes, and error bars represent the median, IQR, and min-max range. The black and grey dashed line indicate the spike-specific IgG MFI for Healthy Controls at D35 and Pre-pandemic samples, respectively. HC = healthy controls; PP = pre-pandemic; CO = cut-off; MFI = median fluorescence intensity.

### Evolution of anti-spike IgG in saliva and serum is harmonized and strongly correlated after vaccination

Next, a receiver operating characteristic (ROC) curve analysis was performed to determine the performance of the salivary anti-spike IgG conversion classified by the present assay relative to the clinical serology result. As shown in **Figure 5A**, area under the curve (AUC) scores raised from 0.82 (D10) to 0.93 (D21) and 0.95 (D35) for anti-Spike-f, and 0.73 (D10) to 0.87 (D21) to0.87 (D35) for the anti-S1 responses. This was also assessed at the respective patient group level (**Figure 5B**). Here, we found that AUC scores in PID, HSCT/CAR-T, SOT, and CLL reached 0.92, 0.99, 0.96, and 0.90 respectively, for anti-Spike-f and 0.87, 0.99, 0.89, and 0.90 for the anti-S1 antibodies, respectively. Due to the very high rates of seroconversion in the healthy control and HIV groups, they were excluded from the analysis. Because the Spike-f antigen appears superior in detecting seroconverted participants, it was chosen for further evaluations against serology data. Based on the serology adjusted cutoff >50 MFI for Spike-f (**Supplementary figure 4**), the endpoint (D35) saliva antibody result yielded 96.3.8% in sensitivity and 73.8% in specificity, relative to the paired serology data when the entire cohort i.e., all patients and healthy controls, were considered. The D35 data also yielded a positive predictive value (PPV) of 90.7% and negative predictive value (NPP) of 88.2% (**Figure 5 C**). Similarly high levels of performance were seen when PID, HSCT, SOT and CLL groups’ anti-Spike-f data was analyzed separately (**Figure 5C**). Altogether, this result confirms the consistently strong serum-to-saliva correlations seen in **Figures 3 and 4**. These data indicate that saliva is functional and accurate in predicting seroconversion as measured in blood.

**Figure 5:**
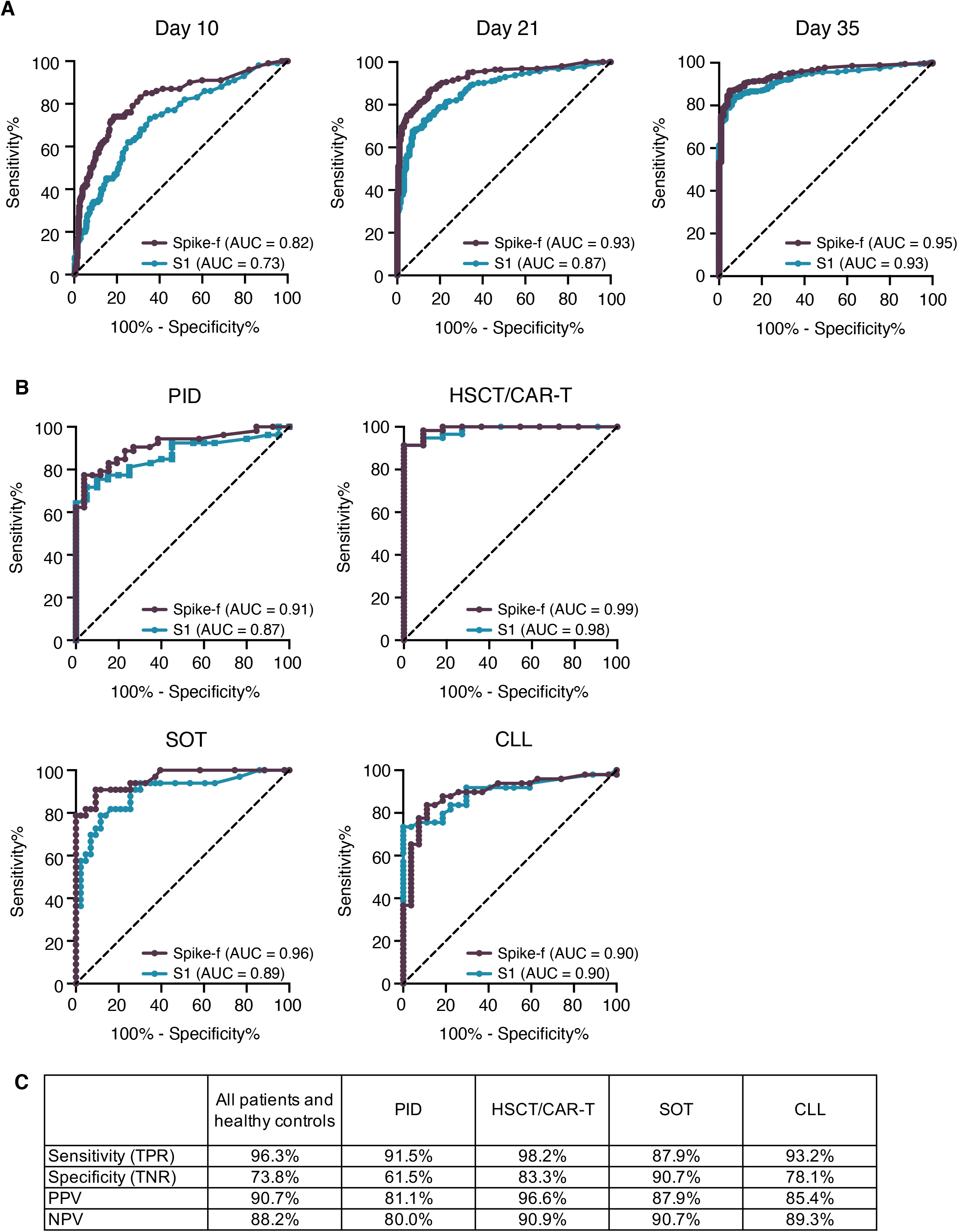
Determination of seroconversion in blood using saliva samples. (A) Graphical representation of the ROC AUC curves for salivary Spike-f and S1 IgG reactivities for the entire study cohort on indicated day after initial vaccination, using the clinical IgG serology result determined by Elecsys® Anti-SARS-CoV-2 S test as reference. (B) ROC AUC curve analysis of indicated patient groups on D35 saliva samples. (C) Performance summary on D35 salivary anti-Spike-f IgG to detect seroconversion in paired serum classified by the Elecsys® Anti-SARS-CoV-2 S test. ROC = receiver operating characteristic; AUC = area under the curve. PPV= positive predictive value; NPV = negative predictive value.

### Assessment of negative predictors for salivary IgG response after vaccination

Next, we evaluated the risk factors associated with failure of salivary antibody conversion after vaccination, where the anti-Spike-f positivity in the endpoint (D35) saliva samples were considered **(Table 3)**. As shown in the univariable analysis, age and sex had little impact on the salivary IgG response. However, the strongest risk for a failure of salivary anti-Spike-f IgG conversion was found in the SOT patients (OR, 32.14; p<0.001), followed by CLL (OR, 17.94; p<0.001), PID (OR, 13.65; p<0.001), and lastly HSCT patients (OR, 5.19; p<0.01). The exact same rank order for these groups was found for serum regarding the risk for failure of seroconversion (28). Within the patient groups, the attributable risk factors among disease- or treatment-specific parameters were also assessed. Notably, being in an early phase post-HSCT transplantation was a strong negative predictor (OR; 19.2, p<0.02). The MMF or ibrutinib drug usage which are critical medications for the SOT and CLL patients, respectively, also negatively impacted the salivary response substantially (OR; 16.44, p<0.001, and OR; 24.44, p<0.001). These results were confirmed by the multivariate analysis as summarized in **Table 3**.

**Table 3.** Logistic regression, univariable and multivariable analysis, assessing variables for failure of salivary antibody conversion to Spike-f after 2 doses of BNT162b2 in D35 saliva samples.

|  | Univariate |  | Multivariate |  |
| --- | --- | --- | --- | --- |
| All | p-value | OR (CI) | p-value | OR (CI) |
| Age | **0.04** | 1.01 (1-1.03) | - | - |
| Sex (M/F) | **0.03** | 1.61 (1.06-2.47) | **0.02** | 1.73 (1.08-2.82) |
| Patient groups: |  |  |  |  |
| PID | **<0.001** | 13.65 (4.49-59.49) | **<0.001** | 14.12 (4.62-61.76) |
| HIV | 0.28 | 2.18 (0.55-10.63) | 0.34 | 1.99 (0.5-9.77) |
| HSCT | **0.01** | 5.19 (1.56-23.58) | **0.02** | 4.94 (1.48-22.5) |
| SOT | **<0.001** | 32.14 (10.74-139.33) | **<0.001** | 32.16 (10.7-139.76) |
| CLL | **<0.001** | 17.94 (5.97-77.75) | **<0.001** | 16.26 (5.38-70.69) |
| Healthy | - | Reference | - | Reference |
| **PID** | **p-value** | **OR (CI)** | **p-value** | **OR (CI)** |
| Age | 0.74 | - | - | - |
| Sex (M/F) | **0.03** | 2.91 (1.09-8.03) | - | - |
| Subgroups: |  |  |  |  |
| CD4 cytop. | **0.04** | 0.11 (0.01-0.63) | - | 0.11 (0.01-0.63) |
| Monogenic disease | 0.07 | - | - | - |
| Other | 0.05 | 0.12 (0.01-0.71) | - | 0.12 (0.01-0.71) |
| XLA | 0.99 | - | - | - |
| CVID | - | Reference | - | Reference |
| **HSCT** | **p-value** | **OR (CI)** | **p-value** | **OR (CI)** |
| Age | 0.22 | - | 0.16 | - |
| Sex (M/F) | 0.4 | - | - | - |
| Subgroups: |  |  |  |  |
| Early | **0.02** | 19.2 (1.58-460) | **0.03** | 17.16 (1.39-415) |
| Intermediate | 0.12 | 3.6 (0.64-17.95) | 0.06 | 5.68 (0.9-36.6) |
| Late | - | Reference | - | - |
| **SOT** | **p-value** | **OR (CI)** | **p-value** | **OR (CI)** |
| Age | 0.97 | - | - | - |
| Sex | 0.25 | - | - | - |
| MMF (yes/no) | **<0.001** | 16.44 (5.51-56.53) | **<0.001** | 16.41 (5.32-59.36) |
| **CLL** | **p-value** | **OR (CI)** | **p-value** | **OR (CI)** |
| Age | 0.56 | - | - | - |
| Sex (M/F) | 0.64 | - | - | - |
| Subgroups: |  |  |  |  |
| Ibrutinib | **<0.001** | 24.44 (6.07-132.85) | **<0.001** | 17.13 (4-96.37) |
| Off Ibrutinib | **0.03** | 7.33 (1.21-52.21) | 0.14 | - |
| Prev CD20 mAb therapy | 0.17 | - | 0.28 | - |
| Indolent | - | Reference | - | - |

<sup>a</sup>: Variables with p-value $\leq 0.35$ in univariable analysis were submitted to multivariable analysis. In the multivariable analysis, the variables retained in the final model after stepwise selection procedure are shown (odds ratios and CI are shown only for the significant variables).
<sup>b</sup>: For variables with categories of yes (Y) or no (N), no was set as reference group. Abbreviations: XLA, X-linked agammaglobulinemia; CVID, common variable immunodeficiency; MMF, mycophenolate mofetil, CD20 mAb, cluster of differentiation monoclonal antibody; Off ibrutinib, off ibutinib treatment for >2 months; Indolent, indolent and not treated. OR: odds ratio, CI: 95% confidence interval.

## DISCUSSION

A central clinical question in the global COVID-19 vaccination effort is the effectiveness of the new vaccines in immunocompromised individuals, including how effective a parentally administered new vaccine may provide mucosal immunity in these vulnerable individuals. While most studies focus on the immune markers in blood, this study focused on salivary immunity markers in these individuals. We report here that BNT162b2 vaccine-induced IgG levels in saliva in immunocompromised vaccinees may vary but could also reach levels normally acquired from natural SARS-CoV-2 infection, as recently shown in healthcare workers vaccinated with mRNA vaccine (30). Moreover, we identified risk factors for poor antibody induction in saliva, that pointed out several significant negative predictors such as SOT, CLL, PID and disease related treatment regimens i.e., MMF and ibrutinib, among the conditions studied here. These risk factors interestingly mirror the observations we made recently on a study using serum samples from this cohort (28). Therefore, our results not only demonstrate that the presence of salivary antibodies to the viral spike is strongly connected to the circulating antibodies, also it shows a strong performance in assessing seroconversion in blood that possibly could serve for a diagnostic purpose. To our knowledge, this is the largest SARS-CoV-2 vaccine study using saliva as a biofluid for tracking seroconversion, encompassing four different time points of sample collection on 445 age and sex-matched (baseline seronegative) study participants donating over 1800 paired saliva and serum samples.

The oral compartment is covered by mucosal surfaces which are susceptible to SARS-CoV-2 infection (23). Oral mucosa is endowed with various salivary immunochemical mechanisms to repel foreign intruders and is well-explored by mucosal infection and vaccine experts (23, 31). Primary mechanisms supporting oral mucosa permeability for systemic biomarkers include: (i) passive diffusion, (ii) carrier mediated transport, and (iii) endocytosis/exocytosis where material is actively taken up and excreted by cells via the endocytic pathway (32). Saliva is therefore a functional biofluid and has the potential to mirror systemic antibody responses. Both clinical and experimental data have shown that induction of mucosal immune responses after vaccination might significantly contribute to protection against mucosal infections of respiratory or enteric pathogens (23). Our data further confirms that the mechanism through which serum to saliva transudation occurs appears to operate well in some immunocompromised patients, particularly those living with HIV infection or who have undergone HSCT. In the HIV group, we did not find differences in humoral responses when stratified by low and high CD4 T cell count (at 300 cells/μL). In HSCT patients at least six months after immunocyte reconstitution (the late group), two doses were required to reach the antibody level of healthy controls. Sex and age appeared to have little influence on the salivary antibody responses, which is in-line with serum data reported in earlier cohort studies of HIV and hematological malignancy patients (23). The present data is, to the best of our knowledge, the first to report that swift mucosal antibody responses are in place after COVID-19 mRNA vaccination in certain immunocompromised risk groups.

In patients with PID, SOT or CLL, salivary antibody responses were highly variable. As expected, the XLA and CVID patients were poor antibody responders due to absent (XLA) or impaired (CVID) immunoglobulin production. An Israeli study of 26 PID patients reported seroconversion in the majority of CVID patients, but anti-spike antibody levels remained low relative to healthy controls (20), which is consistent with the data presented here. Importantly, no detectable salivary spike IgG was observed in the XLA patients, which supports that the saliva analysis employed in our study is highly specific. We further report that other vaccinated PID subgroups - monogenic diseases and CD4 T cell cytopenia had detectable saliva antibodies, with some patients responding normally in both saliva and serum. However, the median levels were 10-20-fold lower relative to healthy controls and two vaccine doses appeared insufficient for induction of robust immunity.

Next, we observed that SOT patients with a post-transplantation time of more than 6 months and no MMF treatment were more likely to develop salivary responses after two doses of BNT162b2. This not only mirrors the systemic responses shown in paired serum, but also is in line with a blood serology study by Boyarsky et al. who showed that the use of antimetabolites, including MMF, was persistently associated with poor humoral response in SOT patients after two doses of the BNT162b2 vaccine (33). Adding a third BNT162b2 vaccine dose could be an option to increase the level of protection in SOT patients, especially in those with an initially weak serological response (34), and a non-invasive antibody screening strategy could be helpful to identify this subgroup. Our data further confirms that neither sex or age in any marked way impacted salivary or serum antibody responses in the SOT patients, which is consistent with the latter two studies (34). Furthermore, drugs used in CLL treatment can diminish vaccine responses resulting in very low salivary IgG levels. This was found mainly in active or past ibrutinib (a BTK inhibitor) subgroups, although indolent CLL and previous CD20 mAb based therapy were also impacted to some extent. Whether the variation observed could relate to drug compliance is unknown as no biomarkers exist for validation.

Limitations of our study include the lack of antibody isotype analysis and virus neutralizing capacity. The antibody durability in saliva and exploration of local memory B and T cell immunity remain to be investigated. Vaccines other than BNT162b2 also remain to be compared in similar cohorts. Although our data in saliva in consistent with Sahin et al. (35), that after full vaccination healthy volunteers showed a 10-fold increase in producing spike-binding IgG, the present data suggests that the vast majority of HIV and HSCT patients also displayed similar levels as healthy controls. We did not measure IgA, as it had already been shown that the BNT162b2 vaccine elicits mainly IgG and less IgA in saliva in healthcare workers (30).

Mucosal antibodies specific to SARS-CoV-2 are considered important in reducing transmission potential in vaccinated individuals (36). The magnitude of anti-spike IgG responses in the saliva of vaccinated individuals, which exceeded those seen in mild convalescent individuals, is encouraging as it indicates that vaccination might confer a sterilizing immune response in the oral cavity and thereby lower virus transmission. The observation that vaccine-induced IgG efficiently translocates into saliva, with high predictive values of BNT162b2-induced seroconversion, is beneficial for immune surveys. Many risk groups are vulnerable to SARS-CoV-2 infection and need regular monitoring. Therefore, saliva and home sampling represent a safe and convenient alternative.

Saliva sampling is entirely non-invasive, easy, and can be repeated multiple times. It is therefore ideal for real-time monitoring of frail patient groups that are sensitive to infections. It will be a safe and efficient approach for tracking vaccine immunity to support informed decisions and agreeing on protective strategies for these patients, especially towards a re-opening of society. In this context, saliva is highly suitable for vaccine follow-up studies, can be used for monitoring seroconversion and antibody memory after vaccination. Furthermore, saliva-specific antibody studies also depict the local immunity at a crucial site for the SARS-CoV-2 infection. Swift comparisons of vaccine responses in immunocompromised individuals will improve vaccination strategies and identify those likely to remain at risk for COVID-19 for a revaccination. Our data merits a call to accentuate the diagnostic significance of salivary testing.

## METHODS

### Study participants in the clinical trial

We conducted a prospective, open-label clinical trial of BNT162b2 (Comirnaty^®^, Pfizer/BioNTech) with two doses given to immunocompromised patients and healthy controls at Karolinska University Hospital, Sweden. Evaluated in the study was safety and efficacy (28). The two doses of vaccine were given 21 days apart. Immunocompromised patients (n=449) had either PID (n=90), or SID due to infection with HIV infection (n=90), allogeneic hematopoietic stem cell transplantation (HSCT)/chimeric antigen receptor T (CAR-T) cell therapy (n=90), solid organ transplantation (SOT) (n=89), or chronic lymphocytic leukemia (CLL) (n=90). Healthy controls were age and sex matched (n=90). The number of available saliva samples in each patient group was 79 in PID group, 80 in HIV group, 74 in HSCT/CART-T group, 83 in SOT group and 88 in CLL group. Eligible were individuals ≥ 18 years of age, with no known history of SARS-CoV-2 infection. The study was approved by the Swedish Medical Product Agency (ID 5.1-2021-5881) and the Swedish Ethical Review Authority (ID 2021-00451 and 2020-06381). All participants provided written informed consent. The sponsor of the study was Karolinska University Hospital. This trial was registered at EudraCT (no. 2021-000175-37), and clinicaltrials.gov (no. 2021-000175-37). A description of the current trial with protocol is available via SciLifeLab Data Repository with the following doi: 10.17044/scilifelab.15059364 (English version) and 10.17044/scilifelab.15059355 (Swedish version).

As indicated in the flow chart, a total of 486 patients’ saliva were included from the clinical vaccine study, with 445 paired serum samples for the D35 endpoint analysis. As positive controls, samples donated by COVID-19 convalescent patients were used. These patients were SARS-CoV-2 infected during February to March 2020 with mild (n=21) or severe (n=10) COVID-19. They were recruited from a post-COVID-19 follow-up study at Karolinska University Hospital and sampled 3-9 months after infection (mean: 7.03 months). Negative controls were pre-pandemic saliva samples (n=41) collected during 2016-2018. All participants provided written informed consent.

### Sample collection and SARS-CoV-2 antibody detection in saliva

All saliva samples were processed by a standardized protocol in the same laboratory. Briefly, unstimulated whole saliva was self-collected by fasted study participants as described earlier using standardized picture instructions (37). Participants were instructed to passively drool into a clean cup for five minutes after which the saliva was aliquoted in tubes using a transfer pipette. Samples were either submitted at the study site or mailed in by overnight post. All samples were immediately placed at 4°C upon arrival thereafter stored at -80°C on the same day. Prior to antibody analysis, saliva samples were thawed at 4°C and centrifuged at 400 xg for one min at 4°C to separate any debris. The supernatant was transferred to 96-well PCR plates (100 μL/well) and sealed using qPCR foil seals. Inactivation was then performed at 56°C for 30 min in plate format using a thermal cycler and cooled immediately to 4°C before transferring to - 20°C for antibody analysis.

Antibodies binding to the full-length spike glycoprotein in trimeric form (Spike-f) and the S1 subunit were measured by means of a multiplex bead-based assay in the 384-well plate format (38, 39) as previously described. Briefly, the antigens were immobilized on the surface of uniquely color-coded bead identities (IDs) (MagPlex-C, Luminex corp.), and the IDs pooled to generate the bead-array. Saliva samples were diluted 1:5 in assay buffer (39) and incubated with the array. After cross-linking of the antibody-antigen complexes, a R-phycoerithryne-conjugated anti-human IgG antibody (H10104, Invitrogen) was applied for detection of IgG bound to spike. The assay readout was performed using a FlexMap3D instrument and the Luminex xPONENT software (Luminex Corp.). Each assay run included the same set of 12 negative and 4 positive saliva controls. The negative controls were selected among pre-pandemic saliva samples as representative of the background distribution and therefore used to calculate the antigen and assay specific cutoff, allowing to account for inter-assay variability. The positive controls were selected among convalescent samples with mild disease showing clear reactivity to spike. The inter-assay variability, evaluated as the % CV of the 16 control samples included in each assay run, was 10.8% for Spike-f and 12% for Spike S1 on average.

### Sample collection and SARS-CoV-2 antibody detection in serum

Serum samples were analyzed for detection of antibodies to SARS-CoV-2 spike protein receptor binding domain (RBD), using the quantitative Elecsys^®^ Anti-SARS-CoV-2 S test (Roche Diagnostics) (29) on the Cobas 8000 e801pro. The measuring range is between 0.40 to 250 U/mL, and the cut-off value for positive results is ≥ 0.80 U/mL Positive samples with antibody titers of >250 U/mL were re-tested following 1/10 dilution, and in some cases 1/100 dilution with the upper level of measuring range 25,000 U/mL.

### Data analysis and statistics

The salivary antibody data were acquired as median fluorescence intensities (MFI) for each sample and antigen. The antigen and assay specific cutoff for positivity was calculated as the mean plus 6x standard deviation (SD) of the intensity signals of the 12 selected negative controls. The inter-assay variability was estimated for Spike-f and S1 as the average percent CV of the 16 control samples included in all 6 assay runs required to test the samples included in the current study. Statistical analysis was performed using R and R studio (40) for correlation analyses and logistic regression analyses and Prism software v.9 (Graphpad) for all other comparisons. Datasets initially underwent a data normality distribution test. Differences between groups of samples were analyzed by Mann-Whitney U test for univariate analysis. Correlations were determined using Spearman rank correlation. Logistic regression, univariable or multivariable, was used to analyze possible negative predictive factors. P values <0.05 were considered statistically significant. Two-sided p values <0.05 were considered significant.

## Data Availability

The full clinical study protocol is available via the SciLifeLab Data Repository (English version: doi:10.17044/scilifelab.15059364; Swedish version doi: 10.17044/scilifelab.15059355). Anonymous data displayed in the manuscript will be made available upon request to the corresponding author following publication of the present article. Data will be made available in a form not deviating from what is accepted by local regulatory authorities with respect to handling of patient data, and in adherence of the policies of the Karolinska University Hospital and Karolinska Institutet.

## ACKNOWLEDGEMENTS

We thank all the participants enrolled in this study, and all the research and clinical staff at Karolinska University Hospital, especially research/clinical nurses Sonja Sönnert Husa, Kirsti Niemalä, Begüm Eker, Eva Martell, Helena Pettersson, Charlotta Hausmann, Maria Abramsson, Ruza Milosavljevic, Karin Fransson, Karin Linderståhl, Linn Wursé, Cecilia Lång, Anna Löwhagen Welander, Susanne Hansen, Douglas Carrick, Katarina Stigsäter, Susanne Cederberg, Annika Olsson, Ingrid Andrén, and Margareta Gustafsson. For assistance with biobanking, we would like to thank Agne Kvedaraite, Nazila Samimi, Rosita Mario, and Daniela Sofia Ambrosio. We thank Prof. Jan Albert from Clinical Microbiology, Karolinska University Hospital for fruitful discussions.

---

**Supplementary figure 1:**
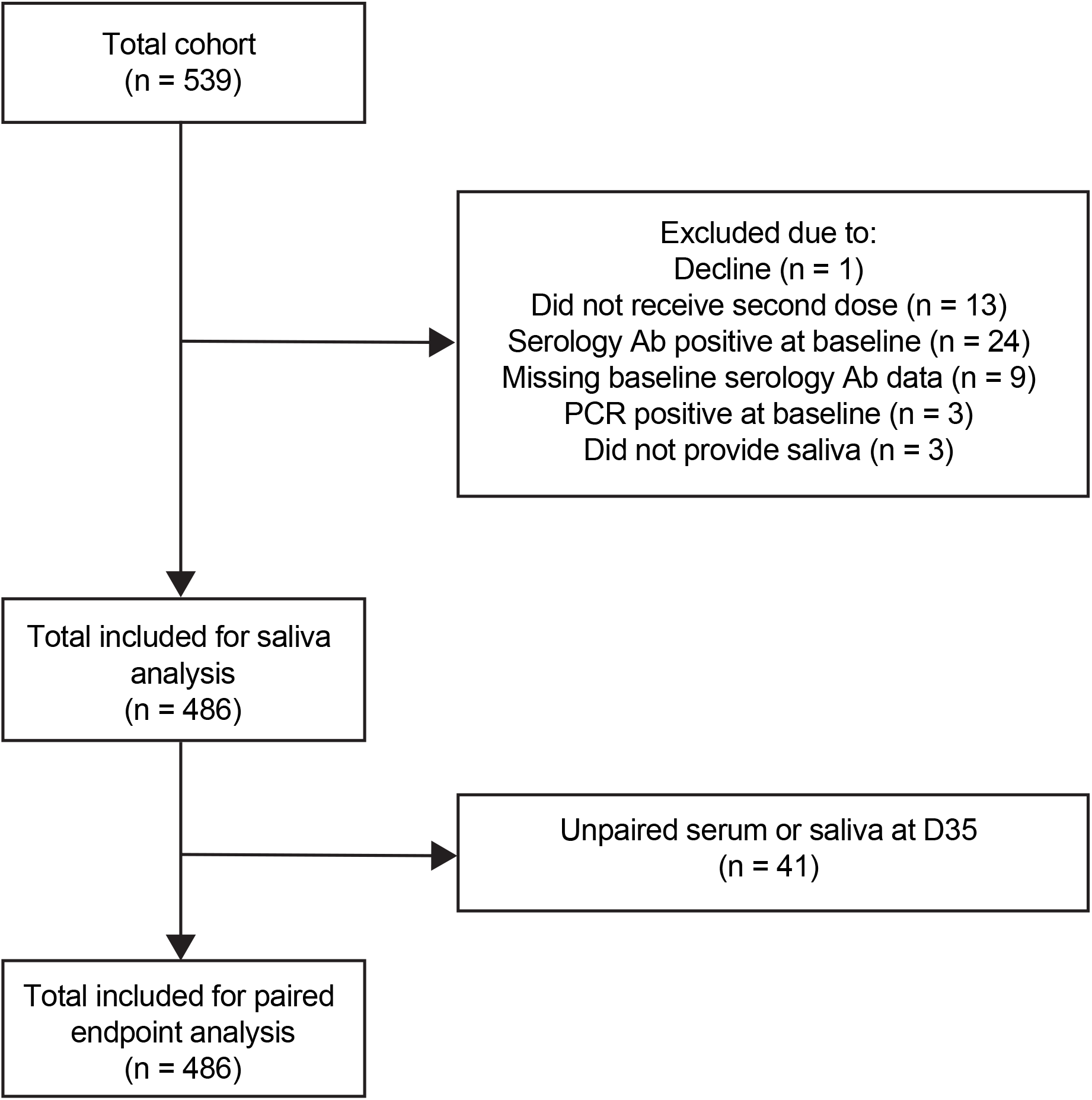
Flow chart of the study design.

**Supplementary figure 2:**
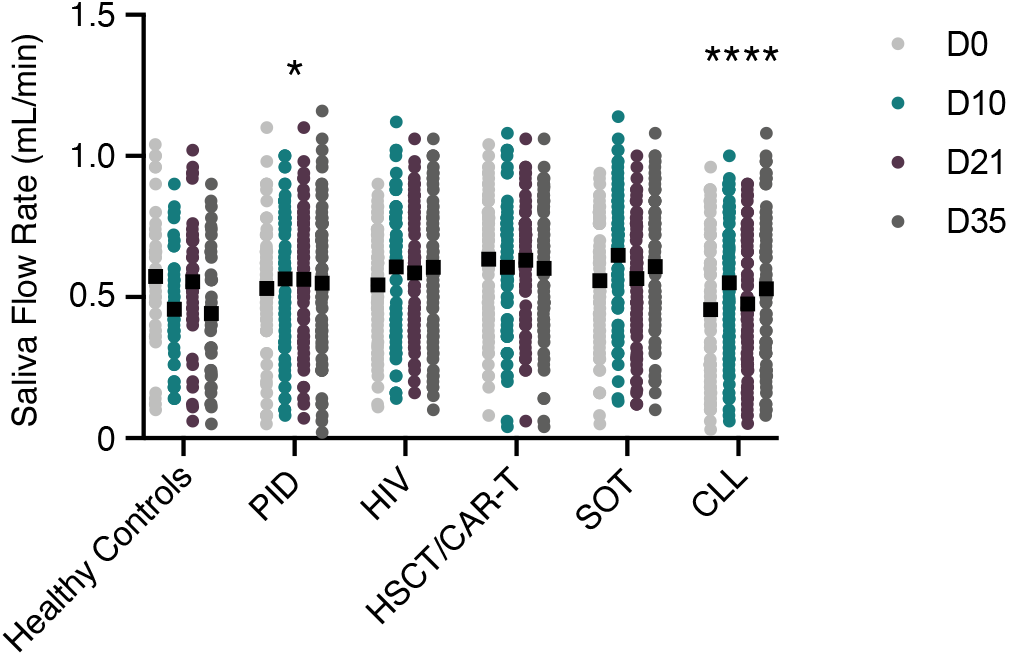
Saliva flow rate for the COVAXID cohort at each timepoint. The Mann-Whitney U test was used to test significance. **** p<0.0001, *** p<0.0002, ** p<0.0021, * p<0.0332. ns = not significant.

**Supplementary figure 3:**
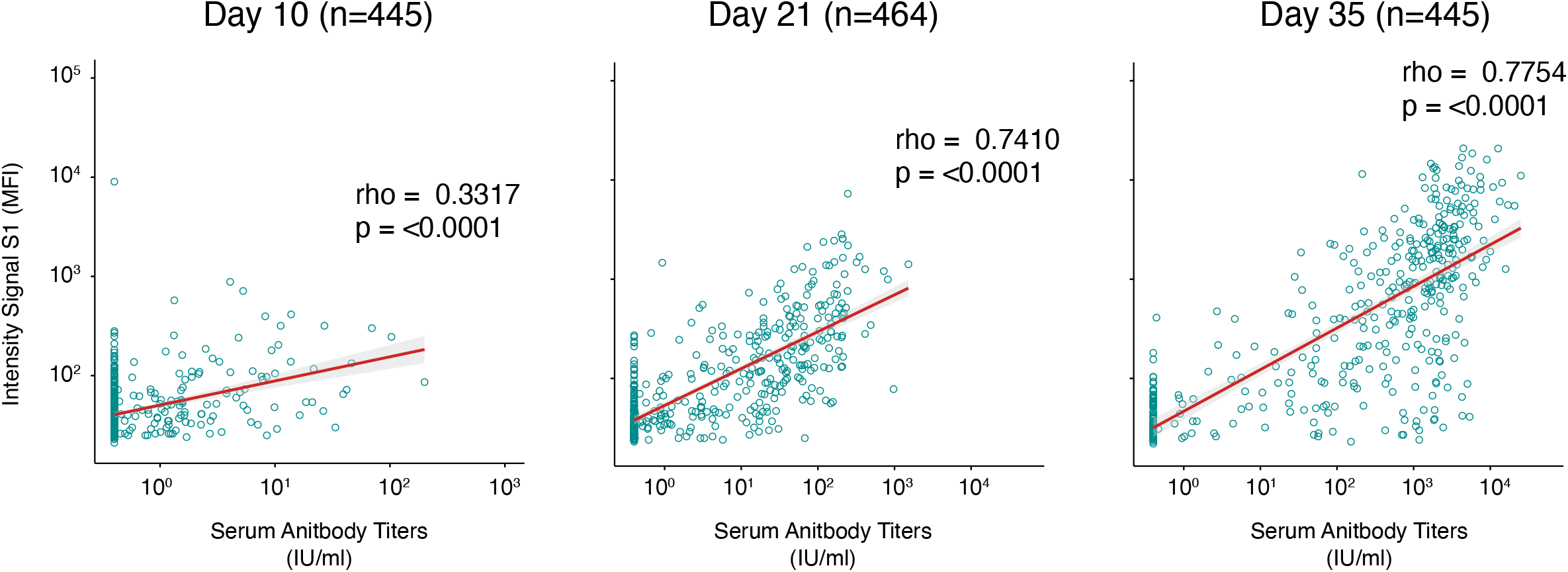
Correlation between spike IgG in paired serum and saliva. Salivary S1 IgG MFI signal intensity (y-axis) was measured by a multiplex bead-based assay, and serum SARS-CoV-2 spike IgG levels expressed as international units (x-axis) were measured by the quantitative test Elecsys® Anti-SARS-CoV-2 S. Correlation plots of the entire COVAXID cohort at D10, D21, and D35 post-vaccination. MFI = Median Fluorescence Intensity; IU = International Units. Spearman correlation analysis was used to determine rho- and p-values.

**Supplementary figure 4:**
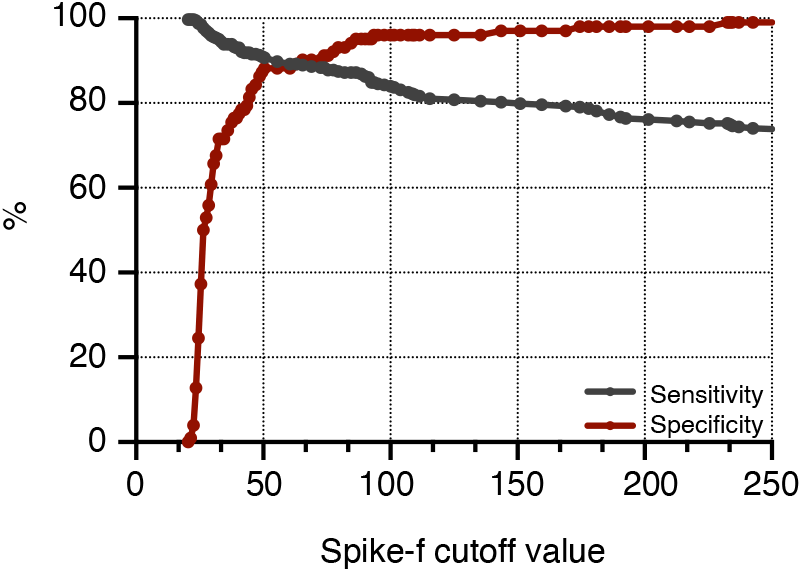
Sensitivity and specificity plot of salivary SARS-CoV-2 Spike-f responses based on optimal cut-off calculation.

## Notes

**DECLARATION OF INTERESTS** The authors have declared that no conflict of interest exists.

### Competing Interest Statement

The authors have declared no competing interest.

### Clinical Trial

NCT04780659

### Funding Statement

This work is supported by research grants from Knut and Alice Wallenberg Foundation, Erling Perssons family foundation, Region Stockholm, Swedish Research Council, Karolinska Institutet, The Swedish Blood Cancer Foundation and the organization for PID patient group in Sweden, and Nordstjernan AB. Center for Medical Innovation (CIMED), the Swedish Medical Research Council and the Stockholm County Council (ALF).

### Author Declarations

The study was approved by the Swedish Medical Product Agency (ID 5.1-2021-5881) and the Swedish Ethical Review Authority (ID 2021-00451 and 2020-06381).

